# Using machine learning to improve diagnostic assessment of ASD in the light of specific differential diagnosis

**DOI:** 10.1101/2021.10.27.21265329

**Authors:** Martin Schulte-Rüther, Tomas Kulvicius, Sanna Stroth, Veit Roessner, Peter Marschik, Inge Kamp-Becker, Luise Poustka

## Abstract

**Background:** Diagnostic assessment of ASD requires substantial clinical experience and is particular difficult in the context of other disorders with behavioral symptoms in the domain of social interaction and communication. Observation measures such as the Autism Diagnostic Observation Schedule (ADOS) do not take into account such comorbid and differential disorders.

**Method:** We used a well-characterized clinical sample of individuals (n=1262) that had received detailed outpatient evaluation for the presence of an ASD diagnosis (n=481) and covered a range of additional differential or overlapping diagnoses, including anxiety related disorders (ANX, n=100), ADHD (n=440), and conduct disorder (CD, n=192). We focused on ADOS module 3, covering the age range with particular high prevalence of such differential diagnoses. We used machine learning (ML) and trained random forest models on ADOS single item scores to predict a clinical best estimate diagnosis of ASD in the context of these differential diagnoses (ASD vs. ANX, ASD vs. ADHD, ASD vs. CD) and an unspecific model using all available data. We employed nested cross-validation for an unbiased estimate of classification performance (ASD vs. non-ASD).

**Results:** We obtained very good overall sensitivity (0.89-0.94) and specificity (0.87-0.89) for the classification of ASD vs. non-ASD. In particular for individuals with less severe symptoms (around the ADOS cut-off) our models showed increases of up to 20% in sensitivity or specificity. Furthermore, we analyzed item importance profiles of the ANX-, ADHD- and CD-models in comparison to the unspecific model. These analyses revealed distinct patterns of importance for specific ADOS-items with respect to differential diagnoses.

**Conclusion:** Using ML-based diagnostic classification may improve clinical decisions by utilizing the full range of information from comprehensive and detailed diagnostic observation such as the ADOS. Importantly, this strategy might be of particular relevance for individuals with less severe symptoms that typically present a very difficult decision for the clinician.

## Introduction

Autism spectrum disorder (ASD) is an umbrella term for a set of highly heterogeneous neurodevelopmental conditions. ASD is characterized by specific impairments in social interaction and communication and restricted, repetitive behaviors (American Psychiatric Association, 2013). The estimated prevalence is ∼1% of the population (Baxter et al., 2015), and there is a considerable amount of overlap with other disorders affecting social interaction (Gjevik et al., 2011; Hossain et al., 2020; Simonoff et al., 2008; Thom et al., 2020). These characteristics call for efficient yet comprehensive and detailed diagnostic procedures. Although ASD is of neurobiological etiology, the diagnosis of ASD is based exclusively on observable behavioral symptoms.

Diagnosing ASD is a challenging, time-consuming task and requires a considerable amount of clinical expertise and experience. The combination of behavioral observation, anamnestic interviews (e.g. Autism Diagnostic Interview – Revised, ADI-R, (Lord et al., 1994)), and additional clinical information (e.g., comorbidity, evaluation of differential diagnoses, cognitive abilities, and neuropsychological impairment) is considered the diagnostic “gold standard.” The Autism Diagnostic Observation Schedule (ADOS-G, ADOS-2, (Lord et al., 2000, 2012; Poustka et al., 2015; Rühl et al., 2004)) is typically used to assess current behavioral symptoms via a structured social interactive encounter with an experienced clinician. It comprises five age-adapted modules (from toddlers to young adults), coding schemes, and diagnostic algorithms providing a simple cut-off score for ASD. However, the actual diagnosis should always be a best-estimate clinical diagnosis, collecting all available information and weighing against other potential differential diagnoses. The direct observation of the respective individual is of utmost importance for this process not only to asses ASD-specific symptomatology, but also to evaluate the behavior in the light of potential differential or comorbid diagnoses.

Diagnostic decisions in ASD are particularly challenging because difficulties in social interaction and communication are common also for a range of other conditions and behaviors including, for example, affective and anxiety disorders (Tyson & Cruess, 2012; van Steensel et al., 2013; Wittkopf et al., 2021), attention deficit hyperactivity disorder (ADHD, Ros & Graziano, 2018; Willis et al., 2019), and conduct disorder (CD, Gilmour et al., 2004; Milledge et al., 2019). Furthermore, the prevalence of respective comorbid disorders among individuals with ASD is high (Hossain et al., 2020). Thus, it is often difficult to decide whether the symptom profile for a specific individual is due to ASD, a differential disorder, or the overlap of both. This decision is particularly difficult for children with less severe ASD symptoms (Davidovitch et al., 2015).

Recent work has explored the potential of machine learning (ML) methods to support diagnostic procedures (see Hyde et al., 2019 for a review). Most of these studies aimed at streamlining the diagnostic procedures for ASD by identifying a reduced set of most discriminative items from clinician-coded examinations such as ADOS-2 and ADI-R (Bone et al., 2015, 2016; Duda et al., 2014; Duda, Daniels, et al., 2016; Kamp-Becker, n.d.; Kosmicki et al., 2015; Küpper et al., 2020; Levy et al., 2017; Stroth, n.d.; Wall, Dally, et al., 2012; Wall, Kosmicki, et al., 2012; Wittkopf et al., 2021, see Bone et al., 2015 for a critical review). ML models built upon such selected items often retain or even exceed the diagnostic accuracy of the original ADOS-2 algorithm. Identifying subsets of items can be the first step towards future developments of time-efficient and sensitive screening instruments (Duda, Daniels, et al., 2016; Kamp-Becker et al., 2017; Omar et al., 2019; Thabtah et al., 2019). However, it is an open question whether such shortened procedures would provide enough specificity in the light of differential diagnostic decisions (Bone et al., 2015). Recent prospective studies aimed to validate shortened parent interviews are promising (Duda, Daniels, et al., 2016), but the results for shortened observation-based approaches (Abbas et al., 2018; Fusaro et al., 2014; Tariq et al., 2018) are mixed.

We suggest a complementary approach that aims to exploit the information given by the entire coding scheme of the ADOS to maximize classification accuracy in the light of specific comorbidity patterns and differential diagnosis. In clinical practice, considering further information (e.g. clinical symptoms of anxiety or externalizing behaviors, IQ) is essential for differential diagnosis and needs to be combined with direct structured observation. This strategy yields additional information beyond the ADOS cut-off score: For example, atypical eye contact or less initiative during social interaction can be weighed differently for obviously anxious individuals, whereas talkative behavior or a lack of empathic responding has different implications for individuals with pronounced externalizing behaviors. However, the standard ADOS algorithm does not take such considerations into account. Providing specific diagnostic algorithms for respective contexts of differential diagnosis would be an essential step in clinical practice and likely increase the overall quality of ASD-specific assessment.

Thus, the present study aimed, for the first time, at using ML methods to train optimized models for ASD diagnosis in the light of specific differential diagnoses (i.e. anxiety, ADHD, and CD). Furthermore, we sought to identify those items of the ADOS which are particular important in the context of such specific models. We focused on ADOS module 3 because the age group covered represents a particular challenge: First, individuals seeking ASD-specific diagnostic service at this age tend to have lower ASD symptoms than those with earlier diagnostic visits (Davidovitch et al., 2015), thus, a higher proportion of “high-functioning” individuals. Second, the onset of comorbid diagnoses such as anxiety related disorders, ADHD, and CD is most prevalent at this age range (Hossain et al., 2020), often resulting in decreased specificity of current diagnostic procedures (Kamp-Becker et al., 2018). We tested i) whether our specific models improve sensitivity and specificity of classification in comparison to the ADOS-2 algorithm, and ii) whether this would be particular pronounced for those individuals with less severe symptoms of ASD. Furthermore, iii) we tested whether item importance profiles of the specific models would reveal those ADOS-items which are particular relevant for the differential diagnostic decision of ASD versus anxiety, ADHD, or conduct disorder.

## Materials and methods

### Dataset and preprocessing

#### Data collection

We used item-level data of the Autism Diagnostic Observation Schedule (ADOS-G/ADOS-2) module 3 (Poustka et al., 2015; Rühl et al., 2004), representing a subsample of data from a German data repository (ASD-Net, Kamp-Becker et al., 2017). All participants visited a specialized outpatient clinic for ASD. A clinical best-estimate diagnosis of ASD was either confirmed or excluded following established guidelines of “gold standard” diagnostic evaluation of ASD (AWMF, 2016; Falkmer et al., 2013). Experienced clinicians with continuous ADOS coding experience performed the coding of all items. Across the sample, we used the ADOS-2 algorithm of module 3. The ADOS-2 algorithm for module 3 is a summed score of a subset of items (cut-off for autism spectrum: 7 or higher, cut-off for autism: 9 or higher). In the following analyses, we used the age-adapted calibrated severity scores, to account for potential age effects across the sample (corresponding cut-offs: 4[autism spectrum], 6[autism]).

#### Missing data

Individuals with more than 50% missing data within the database (n=12) were discarded. The remaining sample had 1.01% missing data across all cells. 92.1% of individuals had no missing data at all.

#### Sample description

The dataset comprised 1262 participants (mean age: 10.06 +-2.73SD), of which 481 received an ASD diagnosis. Participants received additional ICD10 F-diagnoses such as ADHD (Attention Deficit Hyperactivity Disorder, n=440), CD (conduct disorder, n=192), and anxiety related disorders (ANX, n=100). Some individuals had not received either of the diagnostic labels ADHD, ANX, or CD but a different ICD-F diagnosis (OTHER, n=253). These diagnoses were partly overlapping with an ASD diagnosis and with each other. Some individuals did not receive a diagnosis of ASD but had no further assessment to secure a differential ICD-F diagnosis (NONE, n=211). For further details on the exact diagnoses and amount of overlap, see Figure 1 and Table 1.

**Table 1.** ICD-10 diagnoses related to ADHD, CD, and ANX within the full sample

| Diagnosis |  | group | n |
| --- | --- | --- | --- |
| F40 | Phobic anxiety disorders | ANX | 7 |
| F41 | Other anxiety disorders | ANX | 4 |
| F42 | Obsessive-compulsive disorder | ANX | 15 |
| F93.[0-2] | Emotional disorders with onset specific to childhood<br>[separation anxiety, social anxiety, phobic anxiety] | ANX | 40 |
| F91 | Conduct disorders | CD | 47 |
| F92 | Mixed disorders of conduct and emotion | CD | 35 |
| F90 | Attention deficit hyperactivity disorder | ADHD | 440 |
|  | mixed |  |  |
| F90.1 | Hyperkinetic conduct disorder | ADHD, CD | 102 |
| F92.8 | Other mixed disorders of conduct and emotions | ANX, CD | 34 |

**Figure 1.**
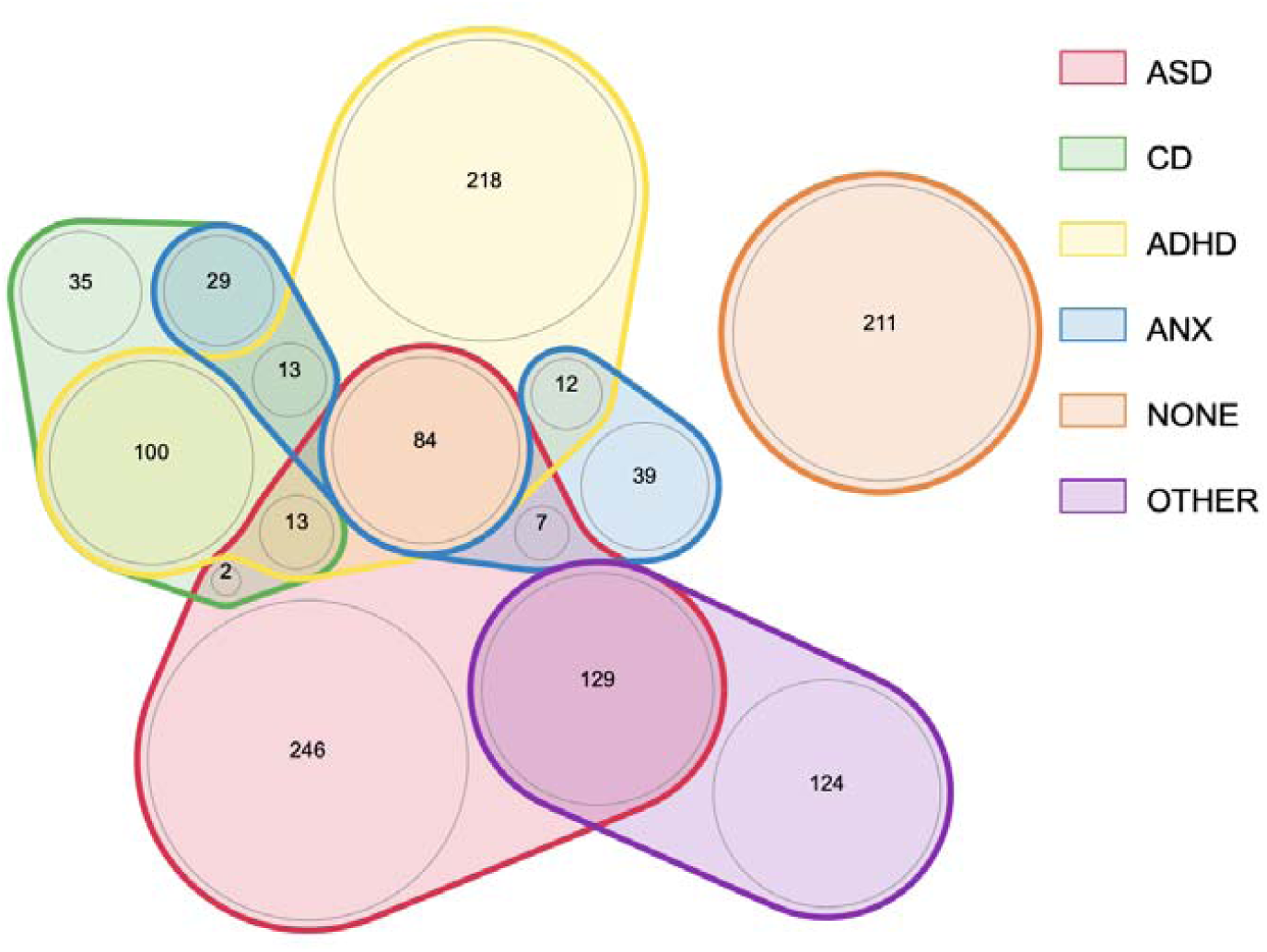
Extent of overlap for diagnostic categories across the sample. For simplicity, the figure only shows overlap across ASD, ADHD, ANX, and CD.

### Classification models

#### Model features

28 items of the ADOS module 3 were used as predictors (item Amount of Social Overtures/ Maintenance of Attention to Examiner of ADOS-2 was excluded because it is only used in ADOS-2 and not in ADOS-G), and the ASD best estimate clinical diagnosis was the target for classification. We used item codes representing an ordinal scale from 0 (no evidence for this type of behavior) to 3 (strong evidence for this type of behavior) and discarded additional categorical codes (e.g. “missing data”, “not applicable”, “other abnormal behavior”).

#### Random forest models

We trained random forest models, as implemented in the ranger package (Wright & Ziegler, 2017), since these provide excellent accuracies for disease prediction from health data when comparing multiple supervised learning algorithms (Uddin et al., 2019). Random forest models typically yield good results with modest parameter tuning and are pretty robust against overfitting. Furthermore, they include a straightforward implementation of permutation-based item importance (Breiman, 2001).

#### Types of models

We used binary classification of ASD vs. non-ASD to train four different sets of models. All models used the full n=481 participants with ASD, but differed with respect to the non-ASD category (ANX-models: Non-ASD[ANX] n= 93, ADHD-models: Non-ASD[ADHD] n=343, CD-models: non-ASD[CD] n= 177, unspecific models: Non-ASD[ANX/ADHD/CD/OTHER/NONE] n= 781). We used this strategy rather than a multilabel classifier, because the ADOS-items are not designed to classify ANX, ADHD, and CD explicitly against each other. Furthermore, we were particularly interested in item importance profiles in the context of specific differential diagnosis. See Table 2 for the characteristics of the ASD and non-ASD samples of each of these models.

**Table 2.** Sample characteristics for the ASD group and each of the non-ASD groups of the respective specific models (ANX, CD, ADHD)

| model | n | Age<br>(mean ± SD) | Wilcoxon-Test (p) | IQ<br>(mean ± SD) | Wilcoxon-Test (p) | no IQ<br>(n) | male | female | $\chi^2$ male<br>/female |
| --- | --- | --- | --- | --- | --- | --- | --- | --- | --- |
| **unspecific**<br>**model** |  |  |  |  |  |  |  |  |  |
| ASD | 481 | 10.3 ± 2.88 | 0.01102 | 99.1 ± 17.0 | 0.4931 | 74 | 438 | 43 | p=0.0595 |
| Non-ASD | 781 | 9.90 ± 2.62 | | 99.6 ± 18.5 | | 204 | 683 | 98 | |
| **CD-models** |  |  |  |  |  |  |  |  |  |
| ASD | 481 | 10.3 ± 2.88 | 0.2313 | 99.1 ± 17.0 | 0.7238 | 74 | 438 | 43 | p=0.5835 |
| Non-ASD | 177 | 9.99 ± 2.43 | | 99.4 ± 20.1 | | 44 | 158 | 19 | |
| **ANX-models** |  |  |  |  |  |  |  |  |  |
| ASD | 481 | 10.3 ± 2.88 | 0.3204 | 99.1 ± 17.0 | 0.286 | 74 | 438 | 43 | p=0.01208 |
| Non-ASD | 93 | 10.6 ± 2.31 | | 101 ± 20.4 | | 23 | 76 | 17 | |
| **ADHD-models** |  |  |  |  |  |  |  |  |  |
| ASD | 481 | 10.3 ± 2.88 | 0.005438 | 99.1 ± 17.0 | 0.8565 | 74 | 438 | 43 | p=0.6779 |
| Non-ASD | 343 | 9.76 ± 2.44 | | 99.2 ± 18.9 | | 80 | 316 | 27 | |

#### Model training and cross-validation

The machine learning pipeline was implemented in tidymodels (Kuhn, Max, 2020) for R (R Core Team, 2020). For a complete list of used modules and versions see the Supplementary Methods. We employed a strict nested cross-validation approach to provide an unbiased estimation of model performance and safeguard against overfitting (Cawley & Talbot, 2010; Vabalas et al., 2019). Although computationally expansive, this strategy results in an unbiased estimation of the model performance and its hyperparameter search.

We used 5-fold repeated cross-validation (r=10) for model evaluation (the outer fold of the nested cross-validation approach), resulting in 50 evaluation models for each type of model (ANX, CD, ADHD, unspecific). Evaluation was performed on 20% of the respective sample, i.e. independent of the 80% training data for each respective model, by calculating area under the curve of the receiver-operant characteristic (AUC), specificity, and sensitivity (subsequently averaged across models). For training, parameters were optimized using five-fold repeated cross-validation (r=5) on just the respective training data for each of the 50 models by splitting these again into test and training sets (the inner fold of the nested cross-validation approach). Bayesian tuning was performed to determine the parameter combinations with the best results (see Supplementary Methods for details). We tuned m_try (number of variables to split at each node), min_n (minimal node size), and trees (number of trees); all other parameters were left at default values. AUC was used as the criterion to evaluate model performance and for optimizing. Subsequently, the mean (across the 25 inner folds) of the respective best combination of parameters for each tuning cycle was used for fitting the respective outer model. We used stratification (based on the outcome variable) to split into training and test sets (inner and outer folds). Removing of zero-variance predictors and imputation of missing values (bagged-tree imputation) was preformed separately for each iteration of parameter optimization and model evaluation to prevent information leakage.

#### Evaluation metrics

For model evaluation, the respective ROC curve for each outer model was used to define optimal cut-points. An iterative algorithm determined the cut-point, which maximized the Youden index (sensitivity+specificity-1), providing an unbiased estimate of model performance in the light of differing ratios of ASD vs. non-ASD across the different models. These optimal cut-points were used to calculate sensitivity, and specificity for each model. To provide an additional evaluation for those individuals who pose the greatest challenge to the clinician, we re-calculated model metrics for individuals with less severe symptoms (i.e. including only those participants with calibrated severity scores of 2-5), but using the same cut-points as derived from the complete sample. Metrics for the Standard ADOS score (for both the total sample and the subset sample) were also computed, averaged across folds, and compared against our new ANX, CD, ADHD, and unspecific models using 2-sample t-tests. For further analyses on model specificity see Supplementary Methods and Figure S3

#### Item importance comparisons between models

Permutation-based item importance gives an estimate of how much the model’s performance (here: AUC-metric) decreases when the item is permuted randomly. It includes the gain in classification accuracy by this specific item and the gain with respect to its interaction with all other items. To compare across models, we normalized the values by dividing the item importance value for a specific item by the sum of importance values across all items, separately for each model. Then, normalized item importance values for each item were aggregated across folds by calculating the median and a rank-based confidence interval. We tested for the specificity of item importance profiles for the specific models [ANX, CD, and ADHD] against the unspecific model using Mann-Whitney Tests for each item.

#### Deployment of final model as an app

For demonstration purposes, we then created four final models (ANX, CD, ADHD, unspecific) using all available data and trained respective random forest models, using 5-fold repeated cross-validation (r=5), Bayesian tuning for parameter optimization, and subsequent averaging of best-performing parameter combinations. We created a web app (https://msrlab.shinyapps.io/ml_beta_app_v3/) that would allow for easy implementation and usage of these models in a clinical setting and can be used for further evaluation of this approach. See Supplementary Methods and Figure S4 for more details.

## Results

### Model evaluation

We observed significantly better performance for the new models in comparison to the ADOS-2 algorithm. In particular, the models performed better for those participants with lower ADOS severity levels (see Figure 2, Supplementary Figure S2 for the respective ROC curves). For all models, AUC was higher for the random forest model than for the ADOS algorithm (all T>5.64, p<.0001, increases of 3-6%). For the full samples (including all severity levels), sensitivity in comparison to the classic ADOS score was higher for the CD (T=6.14, p<0.0001, increase of 5%), and ADHD (all T = 7.32, p< 0.0001, increase of 6%) models, and comparable for the ANX model (T = -0.371, p=0.712, increase of 1%). In contrast, specificity was considerably higher for the ANX model (T=9.96, p<0.0001, increase of ∼12%), and (albeit to a lesser extent) also higher for the CD (T=3.77, p<0.001, increase of 3%) and ADHD models (T=2.48, p<.05, increase of 2%). The random forest models performed considerably better for those individuals with less severity (i.e., ADOS severity levels 2-5) in terms of AUC (T>7.45, p<0.0001, increases of 10-16%), and both with respect to sensitivity (all T >2.84, p<.01) and specificity (all T >3.04, p<.005). The increase in specificity for this subsample was particularly strong for the ANX model (+19%) and less for the CD (+6%) and ADHD models (+5%), whereas the increase in sensitivity was particularly strong for the CD (+20%) and ADHD models(+21%) and less strong for the ANX models (+9%).

**Figure 2.**
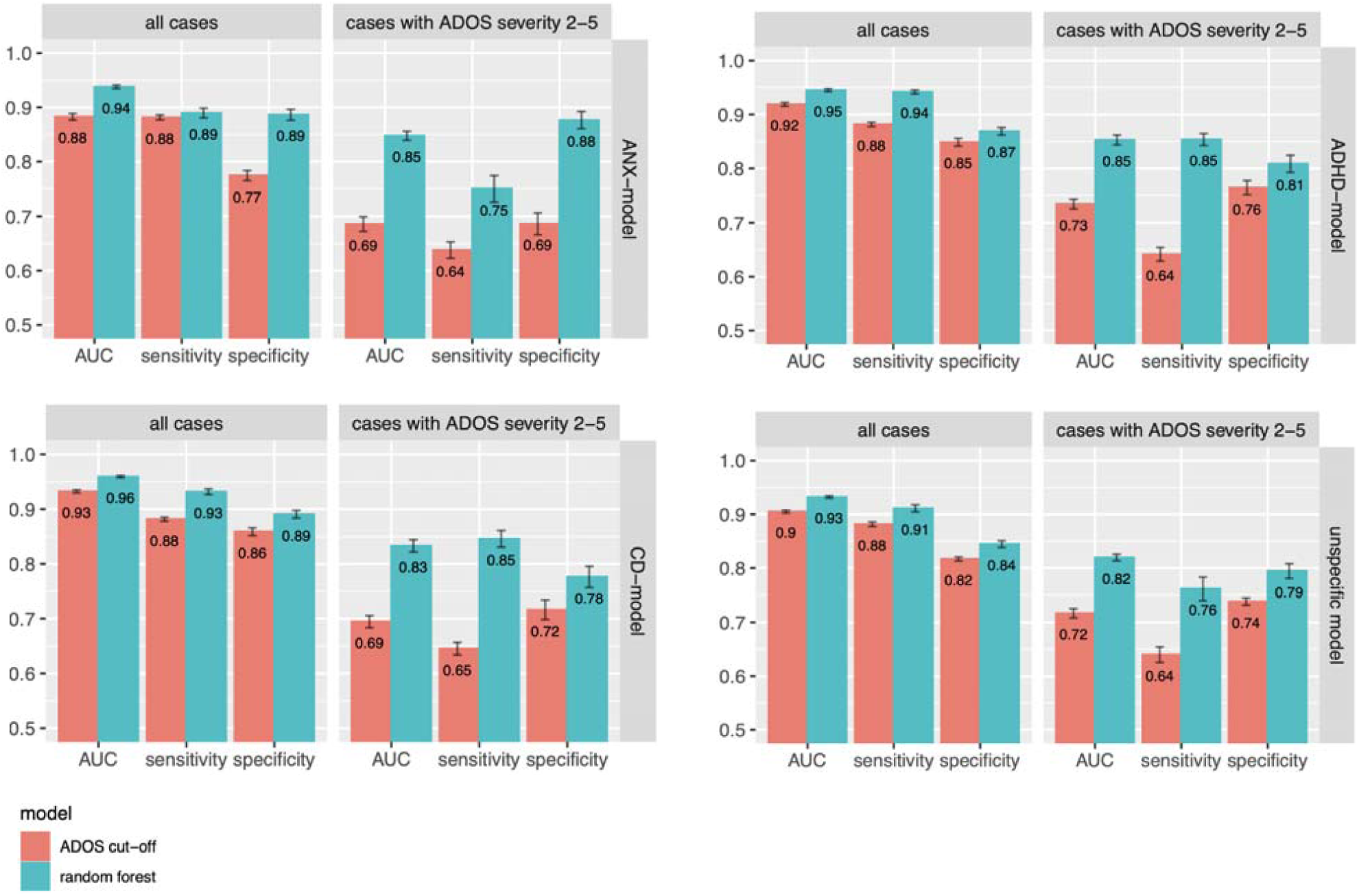
Model evaluation metrics of ANX, CD, ADHD, and unspecific models. Mean AUC, sensitivity and specificity is given for cross-evaluation results and in comparison to the ADOS-2 algorithm, error bars show the standard error of means. Left panels: all available cases, right panels: cases with ADOS severity score 2-5, i.e., around the ADOS cut-off of 4 for an Autism spectrum diagnosis.

### Item importance

Comparing ANX, CD, and ADHD models with the unspecific models for all participants, we could observe unique patterns of item importance (see Figure 3 for profile plots and Figure 4 for difference plots). The item profiles were pretty stable across evaluation model folds, with significant differences for most items (multiple Mann-Whitney tests) in comparison to the profile for the unspecific models (p<0.05 Bonferroni-corrected for multiple comparisons, *CD models*: items SPAB, IECHO, STER, ASK, REPT, DGES, EXPE, LLNC, ENJ, INS, QSOV, QSR, ARSC, OQR, SINT, XINT, ANX; ADHD models: items SPAB, STER, OINF, REPT, CONV, EYE, EXPE, LLNC, INS, QSOV, QSR, ARSC, OQR, IMAG, SINT, RITL, OACT, ANX; ANX model: items SPAB, IECHO, STER, OINF, REPT, CONV, DGES, EYE, EXPE, LLNC, EMO, QSOV, QSR, ARSC, OQR, SINT, MAN, INJ, OACT, TAN, ANX. For a visualization of raw item ratings separately for each diagnostic group see Supplementary Figure S1.

**Figure 3.**
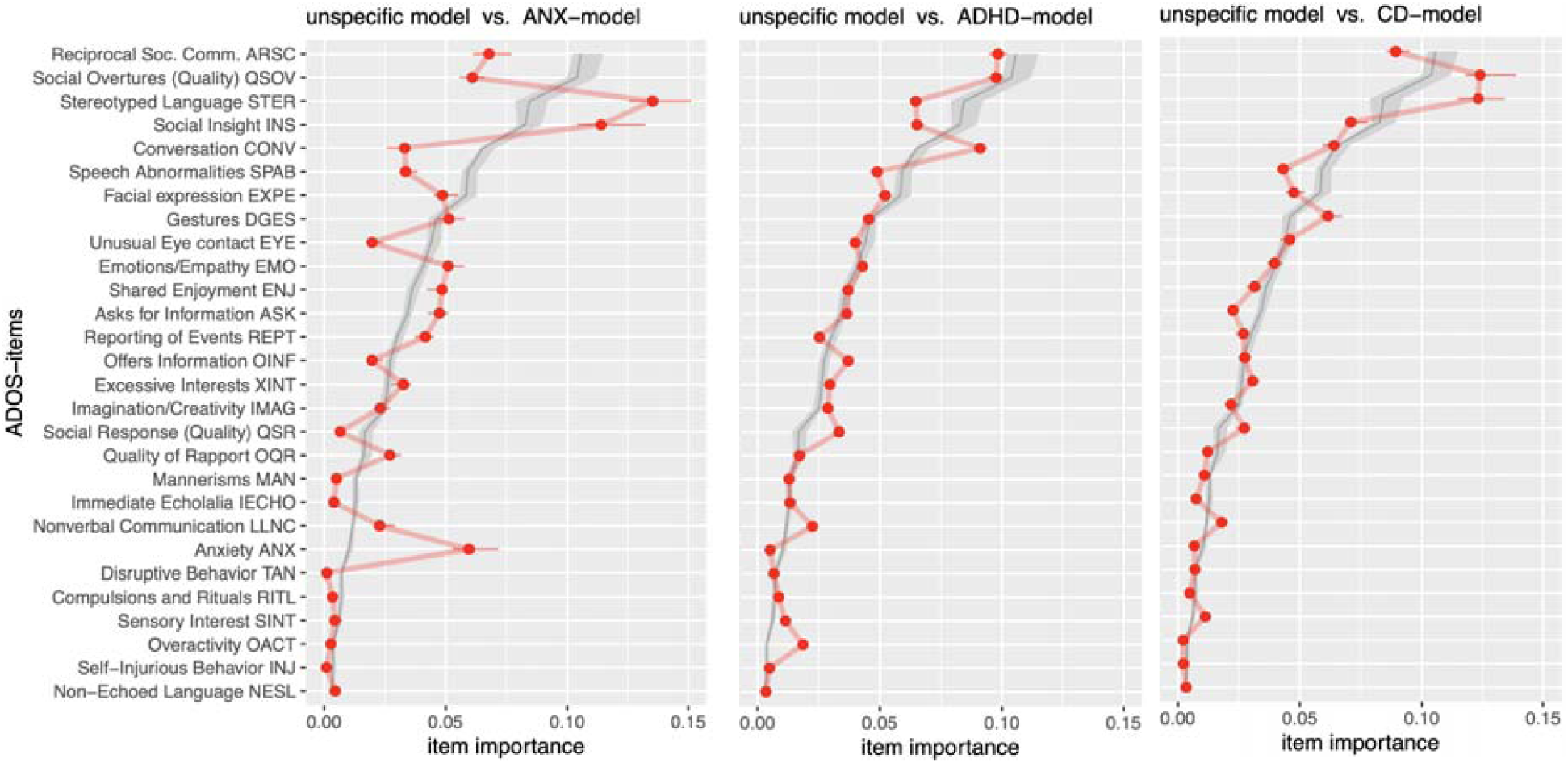
profile plots for ANX, CD, and ADHD models. Red dots and lines indicate the median normalized permutation-based item importance values across evaluation folds for the respective models, with bars indicating a 99% rank-based confidence interval. The black line indicates the median importance values for the unspecific models, with grey shading indicating a 99% rank-based confidence interval. ADOS items are sorted along the y-axis with increasing importance for the unspecific models. Verbal description of ADOS-items are slightly abbreviated, for the full labels refer to Supplementary table S1.

**Figure 4.**
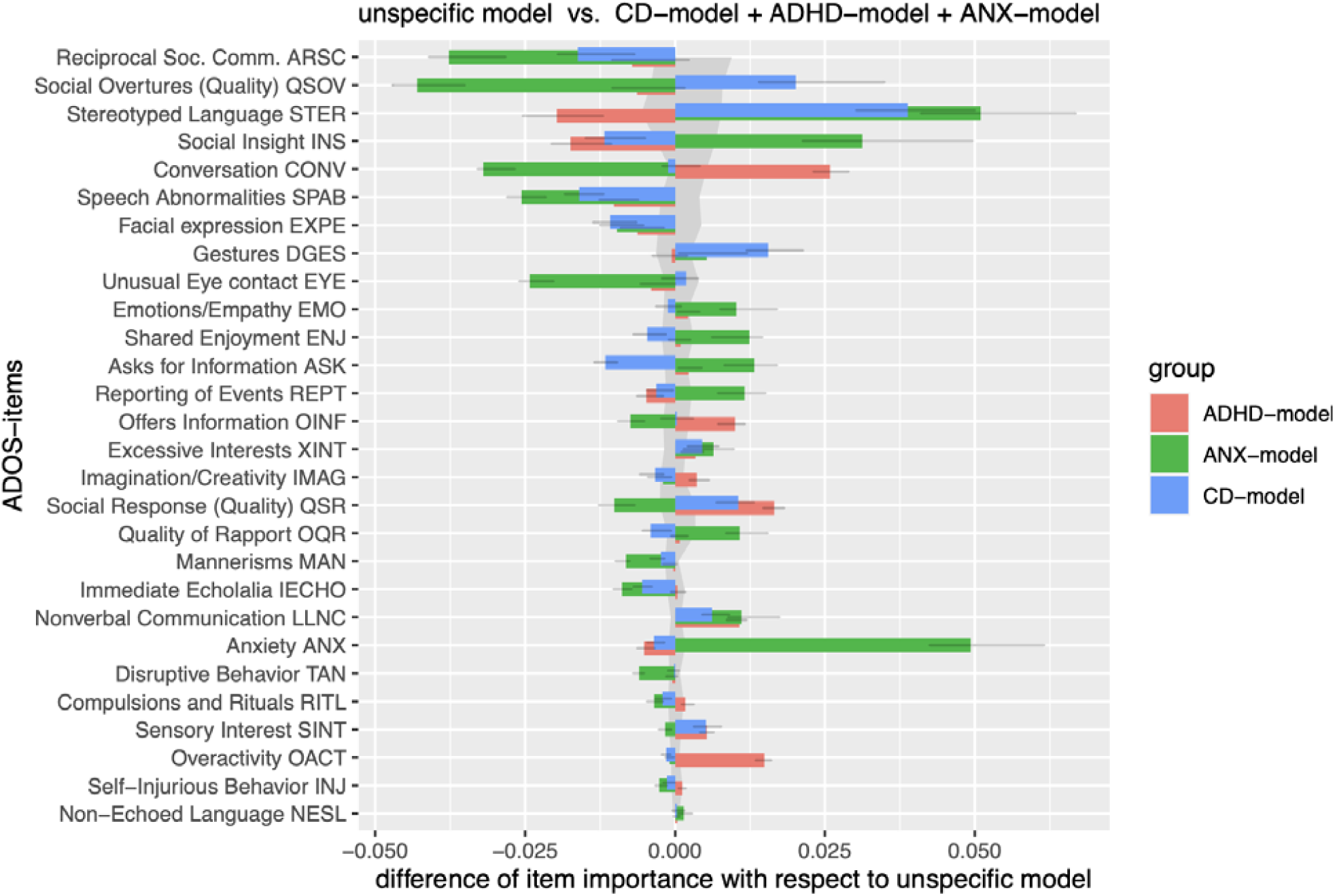
difference plots of normalized item importance for ANX, CD, and ADHD models, with respect to the unspecific models (zero-line). Colored bars indicate the difference to the unspecific models after the respective models’ normalized permutation-based item importance values were determined across evaluation folds. Error bars indicate a 99% rank-based confidence interval of the median. The grey shading indicates a 99% rank-based confidence interval for the unspecific models. ADOS items are sorted along the y-axis with increasing importance for the unspecific models (see also Figure 3). Verbal description of ADOS-items are slightly abbreviated, for the full labels refer to Supplementary table S1.

## Discussion

In this study, we aimed to train optimized ADOS-based models for ASD classification in the light of typical differential diagnoses using machine learning methods. We demonstrate improved sensitivity and specificity in comparison to the ADOS algorithm, in particular for individuals with less severe symptoms. Furthermore, we could reveal specific item importance profiles for the classification of ASD against anxiety related disorders, ADHD, and CD, respectively.

### Improved diagnostic accuracy

#### Model performance

Most previous ML approaches suffer from small, insufficiently characterized samples of non-ASD individuals or a lack of availability of clinical best estimate diagnoses for all individuals (Bone et al., 2015; Duda et al., 2014; Kosmicki et al., 2015; Levy et al., 2017; Wall, Dally, et al., 2012; Wall, Kosmicki, et al., 2012). Within our well-characterized clinical sample, our ML-models showed very good performance to classify ASD (0.94-0.96 AUC, 0.89-0.94 sensitivity, 0.87-0.89 specificity, better than the ADOS-2 algorithm). Such findings indicate that ML is a promising tool to improve existing instruments for ASD diagnosis (Hyde et al., 2019) in a typical clinical population. The resulting models show comparable or better performance in comparison to other studies with much smaller samples of available clinical best-estimate diagnosis (e.g., Duda et al., 2014). Note, other studies reporting higher accuracies typically use the ADOS score and not the clinical diagnosis as their classification target, or a mixture of both. (e.g., Kosmicki et al., 2015; Levy et al., 2017) which may inflate performance measures (Bone et al., 2015).

Previous studies mainly focused on identifying subsets of ADOS-items (Duda et al., 2014; Duda, Daniels, et al., 2016; Duda et al., 2017; Küpper et al., 2020; Wall, Dally, et al., 2012; Wall, Kosmicki, et al., 2012) to streamline the time-consuming diagnostic process. This is a valid and important approach since waiting times for a diagnostic appointment directly translate into delays for the delivery of efficient behavioral therapy. However, the sensitivity and specificity of such shortened instruments need to be ascertained anew since the validity of ADOS-based ratings outside of a complete ADOS examination cannot be assumed (Bone et al., 2015). Furthermore, despite the availability of an efficient screening instrument, detailed, observation-based assessment of the individual is still needed to evaluate the full range of behavioral symptoms. Our results demonstrate that improved algorithms can increase sensitivity and specificity of the ADOS assessment, particularly for children and adolescents who present at diagnostic assessment at an older age. This group is more likely to have less severe ASD symptoms (Davidovitch et al., 2015), often mixed with or masked by comorbid symptoms (Frenette et al., 2013), and respective diagnostic decisions can be challenging. Precisely for these individuals, our approach of optimized models for specific differential diagnoses may better support such difficult clinical decisions.

#### ASD classification in the context of differential diagnoses

Optimized classification algorithms in the context of differential diagnoses can be helpful in clinical practice because other available clinical information (such as clinical anxiety symptoms or externalizing symptoms) often suggests such alternative diagnostic decisions. Respective systematic ML approaches are scarce, to date (but see Duda et al., 2017; Duda, Ma, et al., 2016, for ASD vs. ADHD, and Wittkopf et al., 2021 for ASD vs. mood and anxiety disorders). This might be due to a lack of curated databases of ADOS data which provide at the same time sufficient detail with respect to diagnostic categories other than ASD. The ASD-net (Kamp-Becker et al., 2017) database is unique in this respect by providing a sizeable clinical population with a range of recorded comorbid and differential diagnoses. Here, we could demonstrate, for the first time, the feasibility of constructing optimized models of ASD-classification for specific differential diagnoses. Importantly, the increase in classification performance was particularly strong for individuals that typically present a difficult decision for the clinician, i.e., individuals with lower severity around the ADOS cut-off (up to 20% increase of specificity or sensitivity). Interestingly, the model with the anxiety group showed the largest increase in *specificity* for an ASD diagnosis, whereas the improvement for the models with the CD and ADHD samples was more related to increased *sensitivity*. This finding resonates well with other studies demonstrating lower specificity of the ADOS for Mood and Anxiety disorders (Wittkopf et al., 2021) and a high amount of symptom overlap (Hartley & Sikora, 2009; Tyson & Cruess, 2012; van Steensel et al., 2013). Thus, several individuals with anxiety-related disorders may be misclassified as having ASD when relying too much on the ADOS-cut off scores of module 3. Furthermore, the sensitivity of the ADOS appears to be particularly low for individuals with less severe ASD symptoms in the context of a potential differential diagnosis of CD or ADHD (Grzadzinski et al., 2016; Hartley & Sikora, 2009). Our results demonstrate that optimized classification models can result in substantial improvements of diagnostic accuracy, which may directly translate into improved clinical care.

#### Specific item importance profiles

Our analyses reveal specific item importance profiles of the ADOS with respect to common differential diagnoses of ASD. In comparison to other approaches comparing single items for their potential to distinguish groups (e.g. (Grzadzinski et al., 2011, 2016) for ADHD), these item importance values reflect the total gain of information which may help in distinguishing the diagnostic groups, including non-linear *interactions* between item ratings. Thus, for example, when anxiety is high, a diagnosis of ASD might not always be correct despite high scores on other symptom areas (e.g., eye contact). On the other hand, when hyperactivity is high, excessive talking and missing reciprocity during a conversation might be weighed differently than for less active individuals. In clinical practice, this is precisely what experienced clinicians do when evaluating ADOS results against other behavioral symptoms to arrive at a diagnostic decision (often irrespective of the overall ADOS score). Thus, for example, the increase of the importance value of the anxiety item for the ANX model (5^th^ rank in item importance, in comparison to 23rd rank in the unspecific model) might reflect similar tweaks of the decision trees that compose the respective model and ultimately improve the classification.

#### ANX-model item importance profile

The ANX models had the largest deviations in item importance as compared to the unspecific models. Several items, many of these related to taking the initiative during direct social interaction, such as Amount of Reciprocal Social Communication, Quality of Social Overtures, Conversation, Unusual Eye Contact, were less important for the classification of ASD, along with Speech Abnormalities Associated with Autism and Quality of Social Responses. Other items, such as Stereotyped/Ideosyncratic Use of Words or Phrases, Insight into Typical Social Situations and Relationships, and Anxiety, seemed more important for the classification. To a lesser degree, items such as Comments on Others’ Emotions/Empathy, Shared Enjoyment in Interaction, Reporting of Events, and Asks for Information were also more important for the classification. Interestingly, the item Stereotyped/Ideosyncratic Use of Words or Phrases from the RBB domain was the item with the highest importance for the ANX vs. ASD model (see also (Wittkopf et al., 2021). The RBB domain might be particularly important for the distinction between ASD and anxiety disorders. Other items in this domain (D1-4) were not among those with the highest item importance, but this is in line with the results from the unspecific model and is also compatible with the clinical experience that RBB behavior (as reflected in D-items) is more difficult to observe in ADOS modules 3 and 4.

#### ADHD-model item importance profile

The item importance profile for the ADHD models revealed lower importance for the items Stereotyped/Idiosyncratic Use of Words or Phrases, Insight into Typical Social Situations and Relationships, Anxiety, and Reporting of Events, but higher importance for Conversation, Offers Information, Quality of Social Response, Overactivity, and Language Production and Linked Nonverbal Communication. In contrast, (Grzadzinski et al., 2016) report five items that discriminate best between ADHD and ASD (according to their definition, i.e., that the item is endorsed in >66 % of the ASD group and <33 % of the ADHD group): Quality of Social Overtures, Amount of Reciprocal Social Communication, Unusual Eye Contact, Facial Expressions Directed to Examiner, and Stereotyped/Idiosyncratic Use of Words or Phrases. Although we could replicate this result, i.e. four of these items were among the top 6 items for the ASD vs. ADHD model, our results reveal additionally that this is not necessarily specific to ADHD. Similar items were among those with the highest importance for the unspecific model. In the specific ADHD model, the importance scores of these items were either in the same range or even lower.

#### CD-model item importance profile

The profile for the CD-models revealed slight decreases in item importance for Insight into Typical Social Situations and Relationships, which is in line with the observation of reduced adherence and understanding of social norms and rules in CD. Furthermore, item importance was lower for Speech Abnormalities Associated With Autism (including prosody), Facial Expression Directed to Examiner, Asks for information, and Amount of Reciprocal Social Communication. This finding may be associated with observations of decreased emotional empathy and disturbed affective responsiveness in some individuals with CD, in particular those individuals with high callous-unemotional traits. (Klapwijk et al., 2016; Schwenck et al., 2011; von Polier et al., 2020). These characteristics may translate into lower differentiability for respective associated items in the ADOS. Slight increases of item importance could be observed for Quality of Social Overtures, Stereotyped/ Idiosyncratic Use of Words or Phrases, Descriptive, Conventional, Instrumental, or Informational Gestures, and Quality of Social Response, potentially suggesting stronger differences in behavioral symptoms related to these items for individuals with CD in comparison to ASD.

To summarize, we could reveal differential item importance profiles for ML-based classification based on the ADOS. Future optimized models that draw on an even larger number of individuals, including more fine-grained differential diagnostic groups, could be beneficial to support clinical decisions and better characterize differential profiles of symptom clusters for each potential differential diagnosis.

#### Limitations

The approach presented here is limited to the classification of ASD vs. non-ASD. Since the model builds on an ASD-specific observation instrument, it is not possible to confirm a differential or comorbid diagnosis of ADHD, an anxiety disorder, or CD using these models. Future studies could extend the approach by incorporating a standardized battery of specific diagnostic instruments for a range of diagnostic categories and test whether differential diagnostic decisions can be enhanced by using, for example, multi-label classification approaches. Furthermore, it could be beneficial to include ADI-R items into the models to further enhance diagnostic accuracy. The models currently only allow for the consideration of one specific comorbid/differential diagnosis (i.e. ANX, ADHD, or CD) to improve the ASD diagnosis. With higher sample sizes it would also be possible to distinguish between groups with multiple comorbid diagnoses and to include further characteristics and potentially confounding factors such as sex and age. Lastly, it should be noted that at this stage the models depend on the quality of the ADOS instrument and require highly trained raters as an intermediate step.

#### Future directions

Technological obstacles often hinder translation of ML methods into clinical practice, i.e., distribution of a trained machine learning model available for general use is more difficult than publishing a simple algorithm based on summed items. To advance clinical care and diagnostic accuracy in the upcoming era of personalized medicine, it is essential not only to create large databases and optimized diagnostic algorithms but also to make these available for use in clinical practice. Our app (https://msrlab.shinyapps.io/ml_beta_app_v3/) demonstrates the potential of ML approaches to advance diagnostic decisions in today’s clinical care for ASD. It is intended for research purposes only and should not be used for diagnostic evaluation but provides the opportunity for distributed data collection to evaluate the usability of this or other ML models for clinical practice.

The advantage of clinician rating scales, as used in the ADOS, is that they represent a condensed clinical evaluation of specific dimensions across a broad range of observable behaviors. However, the validity of ratings depends on the clinical expertise and experience of the observer (Kamp-Becker et al., 2018). More quantifiable indices of behavior derived from an ADOS-like examination would be desirable to allow for a more precise and observer-independent assessment. Future developments should incorporate quantifiable assessments of eye gaze (Chong et al., 2019; Hartz et al., 2021), facial expression (Drimalla et al., 2020), motion (Budman et al., 2019), or even neural assessment during social interaction (Kruppa et al., 2020). However, it is unlikely that machine learning approaches will succeed in directly relating the basic building blocks of interactive social behavior to a clinical diagnosis of a heterogeneous disorder such as ASD. A more holistic approach is necessary(Roessner et al., 2021), including intermediate steps: Future studies could aim to set up generative models that describe how quantifiable behavioral indices translate into clinician symptom ratings, and in a second step, relate these to the diagnostic classification. Similar to advances in neuromodeling (Frässle et al., 2018) this approach could use the powerful technique of generative embedding (Shawe-Taylor & Cristianini, 2004) to improve the diagnostic algorithm. Our approach is compatible with such considerations and may encourage further research into this direction.

To conclude, using ML in diagnostic procedures could be an excellent strategy for improving clinical decisions by utilizing the full range of information from comprehensive and detailed diagnostic observation.

## Data Availability

All data produced in the present study are available upon reasonable request to the authors

## Acknowledgements

The authors would like to thank families who participated in the study. In addition, they want to thank Gerti Gerber for support in data management as well as Charlotte Küpper for their cooperation in the development of the data base.

## Data availability

The data and source code for the analyses is available upon reasonable request to the authors.

## Key points

- The diagnosis of autism spectrum disorder in children is particularly challenging in the context of other disorders with behavioral symptoms affecting social interaction. Observation instruments (e.g. ADOS-2) do not take into account such comorbid and differential disorders.
- We trained specific machine learning models based on ADOS-2 items for the classification of ASD versus other disorders.
- We found increased diagnostic accuracy for our specific models, in particular for those patients with less severe ASD symptoms for whom the clinical diagnostic decision is particularly difficult.
- Optimized diagnostic classifier may improve clinical decisions by utilizing the full range of information from comprehensive and detailed diagnostic observation and provide insights into those behavioral symptoms which are particular relevant for differential diagnostic decisions.

## References

Abbas, H., Garberson, F., Glover, E., & Wall, D. P. (2018). Machine learning approach for early detection of autism by combining questionnaire and home video screening. Journal of the American Medical Informatics Association, 25(8), 1000–1007. https://doi.org/10.1093/jamia/ocy039

American Psychiatric Association. (2013). Diagnostic and statistical manual of mental disorders (DSM-5®). American Psychiatric Pub.

AWMF. (2016). Autismus-Spektrum-Störungen im Kindes-, Jugendund Erwachsenenalter Teil 1: Diagnostik. Interdisziplinäre S3-Leitlinie der DGKJP und der DGPPN sowie der beteiligten Fachgesellschaften, Berufsverbände und Patientenorganisationen.

Baxter, A. J., Brugha, T. S., Erskine, H. E., Scheurer, R. W., Vos, T., & Scott, J. G. (2015). The epidemiology and global burden of autism spectrum disorders. Psychological Medicine, 45(3), 601–613. https://doi.org/10.1017/S003329171400172X

Bone, D., Bishop, S. L., Black, M. P., Goodwin, M. S., Lord, C., & Narayanan, S. S. (2016). Use of machine learning to improve autism screening and diagnostic instruments: Effectiveness, efficiency, and multi-instrument fusion. Journal of Child Psychology and Psychiatry, 57(8), 927–937. https://doi.org/10.1111/jcpp.12559

Bone, D., Goodwin, M. S., Black, M. P., Lee, C.-C., Audhkhasi, K., & Narayanan, S. (2015). Applying Machine Learning to Facilitate Autism Diagnostics: Pitfalls and Promises. Journal of Autism and Developmental Disorders, 45(5), 1121–1136. https://doi.org/10.1007/s10803-014-2268-6

Breiman, L. (2001). Random Forests. Machine Learning, 45(1), 5–32. https://doi.org/10.1023/A:1010933404324

Budman, I., Meiri, G., Ilan, M., Faroy, M., Langer, A., Reboh, D., Michaelovski, A., Flusser, H., Menashe, I., Donchin, O., & Dinstein, I. (2019). Quantifying the social symptoms of autism using motion capture. Scientific Reports, 9(1), 7712. https://doi.org/10.1038/s41598-019-44180-9

Cawley, G. C., & Talbot, N. L. C. (2010). On Over-fitting in Model Selection and Subsequent Selection Bias in Performance Evaluation. Journal of Machine Learning Research, 11, 2080–2107.

Chong, E., Chanda, K., Ye, Z., Southerland, A., Ruiz, N., Jones, R. M., Rozga, A., & Rehg, J. M. (2019). Detecting Gaze Towards Eyes in Natural Social Interactions and its Use in Child Assessment. 1902.00607 [Cs]. http://arxiv.org/abs/1902.00607

Davidovitch, M., Levit-Binnun, N., Golan, D., & Manning-Courtney, P. (2015). Late Diagnosis of Autism Spectrum Disorder After Initial Negative Assessment by a Multidisciplinary Team. Journal of Developmental & Behavioral Pediatrics, 36(4), 227–234. https://doi.org/10.1097/DBP.0000000000000133

Drimalla, H., Scheffer, T., Landwehr, N., Baskow, I., Roepke, S., Behnia, B., & Dziobek, I. (2020). Towards the automatic detection of social biomarkers in autism spectrum disorder: Introducing the simulated interaction task (SIT). Npj Digital Medicine, 3(1). https://doi.org/10.1038/s41746-020-0227-5

Duda, M., Daniels, J., & Wall, D. P. (2016). Clinical Evaluation of a Novel and Mobile Autism Risk Assessment. Journal of Autism and Developmental Disorders, 46(6), 1953–1961. https://doi.org/10.1007/s10803-016-2718-4

Duda, M., Haber, N., Daniels, J., & Wall, D. P. (2017). Crowdsourced validation of a machine-learning classification system for autism and ADHD. Translational Psychiatry, 7(5), e1133–e1133. https://doi.org/10.1038/tp.2017.86

Duda, M., Kosmicki, J. A., & Wall, D. P. (2014). Testing the accuracy of an observation-based classifier for rapid detection of autism risk. Translational Psychiatry, 4(8), e424–e424. https://doi.org/10.1038/tp.2014.65

Duda, M., Ma, R., Haber, N., & Wall, D. P. (2016). Use of machine learning for behavioral distinction of autism and ADHD. Translational Psychiatry, 6(2), e732–e732. https://doi.org/10.1038/tp.2015.221

Falkmer, T., Anderson, K., Falkmer, M., & Horlin, C. (2013). Diagnostic procedures in autism spectrum disorders: A systematic literature review. European Child & Adolescent Psychiatry, 22(6), 329–340. https://doi.org/10.1007/s00787-013-0375-0

Frässle, S., Yao, Y., Schöbi, D., Aponte, E. A., Heinzle, J., & Stephan, K. E. (2018). Generative models for clinical applications in computational psychiatry. WIREs Cognitive Science, 9(3). https://doi.org/10.1002/wcs.1460

Frenette, P., Dodds, L., MacPherson, K., Flowerdew, G., Hennen, B., & Bryson, S. (2013). Factors affecting the age at diagnosis of autism spectrum disorders in Nova Scotia, Canada. Autism, 17(2), 184–195. https://doi.org/10.1177/1362361311413399

Fusaro, V. A., Daniels, J., Duda, M., DeLuca, T. F., D’Angelo, O., Tamburello, J., Maniscalco, J., & Wall, D. P. (2014). The Potential of Accelerating Early Detection of Autism through Content Analysis of YouTube Videos. PLoS ONE, 9(4), e93533. https://doi.org/10.1371/journal.pone.0093533

Gilmour, J., Hill, B., Place, M., & Skuse, D. H. (2004). Social communication deficits in conduct disorder: A clinical and community survey. Journal of Child Psychology and Psychiatry, 45(5), 967–978. https://doi.org/10.1111/j.1469-7610.2004.t01-1-00289.x

Gjevik, E., Eldevik, S., Fjæran-Granum, T., & Sponheim, E. (2011). Kiddie-SADS Reveals High Rates of DSM-IV Disorders in Children and Adolescents with Autism Spectrum Disorders. Journal of Autism and Developmental Disorders, 41(6), 761–769. https://doi.org/10.1007/s10803-010-1095-7

Grzadzinski, R., Di Martino, A., Brady, E., Mairena, M. A., O’Neale, M., Petkova, E., Lord, C., & Castellanos, F. X. (2011). Examining Autistic Traits in Children with ADHD: Does the Autism Spectrum Extend to ADHDã Journal of Autism and Developmental Disorders, 41(9), 1178–1191. https://doi.org/10.1007/s10803-010-1135-3

Grzadzinski, R., Dick, C., Lord, C., & Bishop, S. (2016). Parent-reported and clinician-observed autism spectrum disorder (ASD) symptoms in children with attention deficit/hyperactivity disorder (ADHD): Implications for practice under DSM-5. Molecular Autism, 7(1), 7. https://doi.org/10.1186/s13229-016-0072-1

Hartley, S. L., & Sikora, D. M. (2009). Which DSM-IV-TR criteria best differentiate high-functioning autism spectrum disorder from ADHD and anxiety disorders in older childrenã Autism, 13(5), 485–509. https://doi.org/10.1177/1362361309335717

Hartz, A., Guth, B., Jording, M., Vogeley, K., & Schulte-Rüther, M. (2021). Temporal Behavioral Parameters of On-Going Gaze Encounters in a Virtual Environment. Frontiers in Psychology, 12, 673982. https://doi.org/10.3389/fpsyg.2021.673982

Hossain, M. M., Khan, N., Sultana, A., Ma, P., McKyer, E. L. J., Ahmed, H. U., & Purohit, N. (2020). Prevalence of comorbid psychiatric disorders among people with autism spectrum disorder: An umbrella review of systematic reviews and meta-analyses. Psychiatry Research, 287, 112922. https://doi.org/10.1016/j.psychres.2020.112922

Hyde, K. K., Novack, M. N., LaHaye, N., Parlett-Pelleriti, C., Anden, R., Dixon, D. R., & Linstead, E. (2019). Applications of Supervised Machine Learning in Autism Spectrum Disorder Research: A Review. Review Journal of Autism and Developmental Disorders, 6(2), 128–146. https://doi.org/10.1007/s40489-019-00158-x

Kamp-Becker, I. (n.d.). Is the combination of ADOS and ADI-R necessary to classify ASDã Rethinking the “gold standard” in diagnosing ASD.

Kamp-Becker, I., Albertowski, K., Becker, J., Ghahreman, M., Langmann, A., Mingebach, T., Poustka, L., Weber, L., Schmidt, H., Smidt, J., Stehr, T., Roessner, V., Kucharczyk, K., Wolff, N., & Stroth, S. (2018). Diagnostic accuracy of the ADOS and ADOS-2 in clinical practice. European Child & Adolescent Psychiatry, 27(9), 1193–1207. https://doi.org/10.1007/s00787-018-1143-y

Kamp-Becker, I., Poustka, L., Bachmann, C., Ehrlich, S., Hoffmann, F., Kanske, P., Kirsch, P., Krach, S., Paulus, F. M., Rietschel, M., Roepke, S., Roessner, V., Schad-Hansjosten, T., Singer, T., Stroth, S., Witt, S., & Wermter, A.-K. (2017). Study protocol of the ASD-Net, the German research consortium for the study of Autism Spectrum Disorder across the lifespan: From a better etiological understanding, through valid diagnosis, to more effective health care. BMC Psychiatry, 17(1), 206. https://doi.org/10.1186/s12888-017-1362-7

Klapwijk, E. T., Aghajani, M., Colins, O. F., Marijnissen, G. M., Popma, A., Lang, N. D. J., Wee, N. J. A., & Vermeiren, R. R. J. M. (2016). Different brain responses during empathy in autism spectrum disorders versus conduct disorder and callous-unemotional traits. Journal of Child Psychology and Psychiatry, 57(6), 737–747. https://doi.org/10.1111/jcpp.12498

Kosmicki, J. A., Sochat, V., Duda, M., & Wall, D. P. (2015). Searching for a minimal set of behaviors for autism detection through feature selection-based machine learning. Translational Psychiatry, 5(2), e514–e514. https://doi.org/10.1038/tp.2015.7

Kruppa, J. A., Reindl, V., Gerloff, C., Oberwelland Weiss, E., Prinz, J., Herpertz-Dahlmann, B., Konrad, K., & Schulte-Rüther, M. (2020). Interpersonal Synchrony Special Issue Brain and motor synchrony in children and adolescents with ASD—a fNIRS hyperscanning study. Social Cognitive and Affective Neuroscience. https://doi.org/10.1093/scan/nsaa092

Kuhn, Max. (2020). Tidymodels: A collection of packages for modeling and machine learning using tidyverse principles. https://www.tidymodels.org

Küpper, C., Stroth, S., Wolff, N., Hauck, F., Kliewer, N., Schad-Hansjosten, T., Kamp-Becker, I., Poustka, L., Roessner, V., Schultebraucks, K., & Roepke, S. (2020). Identifying predictive features of autism spectrum disorders in a clinical sample of adolescents and adults using machine learning. Scientific Reports, 10(1), 4805. https://doi.org/10.1038/s41598-020-61607-w

Levy, S., Duda, M., Haber, N., & Wall, D. P. (2017). Sparsifying machine learning models identify stable subsets of predictive features for behavioral detection of autism. Molecular Autism, 8(1), 65. https://doi.org/10.1186/s13229-017-0180-6

Lord, C., Risi, S., Lambrecht, L., Cook, E. H., Leventhal, B. L., DiLavore, P. C., Pickles, A., & Rutter, M. (2000). The Autism Diagnostic Observation Schedule-Generic: A standard measure of social and communication deficits associated with the spectrum of autism. Journal of Autism and Developmental Disorders, 30(3), 205–223.

Lord, C., Rutter, M., DiLavore, P., Risi, S., Gotham, K., Bishop, S., & others. (2012). Autism diagnostic observation schedule–2nd edition (ADOS-2). Los Angeles, CA: Western Psychological Corporation, 284.

Lord, C., Rutter, M., & Le Couteur, A. (1994). Autism Diagnostic Interview-Revised: A revised version of a diagnostic interview for caregivers of individuals with possible pervasive developmental disorders. Journal of Autism and Developmental Disorders, 24(5), 659–685.

Milledge, S. V., Cortese, S., Thompson, M., McEwan, F., Rolt, M., Meyer, B., Sonuga-Barke, E., & Eisenbarth, H. (2019). Peer relationships and prosocial behaviour differences across disruptive behaviours. European Child & Adolescent Psychiatry, 28(6), 781–793. https://doi.org/10.1007/s00787-018-1249-2

Omar, K. S., Mondal, P., Khan, N. S., Rizvi, Md. R. K., & Islam, M. N. (2019). A Machine Learning Approach to Predict Autism Spectrum Disorder. 2019 International Conference on Electrical, Computer and Communication Engineering (ECCE), 1–6. https://doi.org/10.1109/ECACE.2019.8679454

Poustka, L., Rühl, D., Feineis-matthews, S., Bölte, S., Poustka, F., & Hartung, M. (2015). ADOS-2 Diagnostische Beobachtungsskale für Autistische Störungen—2. Verlag Hans Huber, Hogrefe AG.

R Core Team. (2020). R: A Language and Environment for Statistical Computing. R Foundation for Statistical Computing. https://www.R-project.org/

Roessner, V., Rothe, J., Kohls, G., Schomerus, G., Ehrlich, S., & Beste, C. (2021). Taming the chaosã! Using eXplainable Artificial Intelligence (XAI) to tackle the complexity in mental health research. European Child & Adolescent Psychiatry, 30(8), 1143–1146. https://doi.org/10.1007/s00787-021-01836-0

Ros, R., & Graziano, P. A. (2018). Social Functioning in Children With or At Risk for Attention Deficit/Hyperactivity Disorder: A Meta-Analytic Review. Journal of Clinical Child & Adolescent Psychology, 47(2), 213–235. https://doi.org/10.1080/15374416.2016.1266644

Rühl, D., Bölte, S., Feineis-Matthews, S., & Poustka, F. (2004). Diagnostische Beobachtungsskala für Autistische Störungen (ADOS). In Bern, Huber. Huber.

http://scholar.google.com/scholar?hl=en&btnG=Search&q=intitle:Diagnostische+Beobachtungsskala+f%EF%BF%BD%EF%BF%BDr+Autistische+St%EF%BF%BDrungen+(ADOS)#0

Schwenck, C., Mergenthaler, J., Keller, K., Zech, J., Salehi, S., Taurines, R., Romanos, M., Schecklmann, M., Schneider, W., Warnke, A., & Freitag, C. M. (2011). Empathy in children with autism and conduct disorder: Group-specific profiles and developmental aspects. Journal of Child Psychology and Psychiatry, and Allied Disciplines. https://doi.org/10.1111/j.1469-7610.2011.02499.x

Shawe-Taylor, J., & Cristianini, N. (2004). Kernel Methods for Pattern Analysis. University Press.

Simonoff, E., Pickles, A., Charman, T., Chandler, S., Loucas, T., & Baird, G. (2008). Psychiatric Disorders in Children With Autism Spectrum Disorders: Prevalence, Comorbidity, and Associated Factors in a Population-Derived Sample. Journal of the American Academy of Child & Adolescent Psychiatry, 47(8), 921–929. https://doi.org/10.1097/CHI.0b013e318179964f

Stroth, S. (n.d.). Rethinking our best practice in diagnostic evaluations of Young Children with Autism Spectrum Disorder.

Tariq, Q., Daniels, J., Schwartz, J. N., Washington, P., Kalantarian, H., & Wall, D. P. (2018). Mobile detection of autism through machine learning on home video: A development and prospective validation study. PLOS Medicine, 15(11), e1002705. https://doi.org/10.1371/journal.pmed.1002705

Thabtah, F., Abdelhamid, N., & Peebles, D. (2019). A machine learning autism classification based on logistic regression analysis. Health Information Science and Systems, 7(1), 12. https://doi.org/10.1007/s13755-019-0073-5

Thom, R. P., Keary, C. J., Kramer, G., Nowinski, L. A., & McDougle, C. J. (2020). Psychiatric Assessment of Social Impairment Across the Lifespan. Harvard Review of Psychiatry, 28(3), 159–178. https://doi.org/10.1097/HRP.0000000000000257

Tyson, K. E., & Cruess, D. G. (2012). Differentiating High-Functioning Autism and Social Phobia. Journal of Autism and Developmental Disorders, 42(7), 1477–1490. https://doi.org/10.1007/s10803-011-1386-7

Uddin, S., Khan, A., Hossain, M. E., & Moni, M. A. (2019). Comparing different supervised machine learning algorithms for disease prediction. BMC Medical Informatics and Decision Making, 19(1), 281. https://doi.org/10.1186/s12911-019-1004-8

Vabalas, A., Gowen, E., Poliakoff, E., & Casson, A. J. (2019). Machine learning algorithm validation with a limited sample size. PLOS ONE, 14(11), e0224365. https://doi.org/10.1371/journal.pone.0224365

van Steensel, F. J. A., Bögels, S. M., & Wood, J. J. (2013). Autism Spectrum Traits in Children with Anxiety Disorders. Journal of Autism and Developmental Disorders, 43(2), 361–370. https://doi.org/10.1007/s10803-012-1575-z

von Polier, G. G., Greimel, E., Konrad, K., Großheinrich, N., Kohls, G., Vloet, T. D., Herpertz-Dahlmann, B., & Schulte-Rüther, M. (2020). Neural Correlates of Empathy in Boys With Early Onset Conduct Disorder. Frontiers in Psychiatry, 11. https://doi.org/10.3389/fpsyt.2020.00178

Wall, D. P., Dally, R., Luyster, R., Jung, J.-Y., & Deluca, T. F. (2012). Use of artificial intelligence to shorten the behavioral diagnosis of autism. PloS One, 7(8), e43855. https://doi.org/10.1371/journal.pone.0043855

Wall, D. P., Kosmicki, J., Deluca, T. F., Harstad, E., & Fusaro, V. a. (2012). Use of machine learning to shorten observation-based screening and diagnosis of autism. Translational Psychiatry, 2(4), e100. https://doi.org/10.1038/tp.2012.10

Willis, D., Siceloff, E. R., Morse, M., Neger, E., & Flory, K. (2019). Stand-Alone Social Skills Training for Youth with ADHD: A Systematic Review. Clinical Child and Family Psychology Review, 22(3), 348–366. https://doi.org/10.1007/s10567-019-00291-3

Wittkopf, S., Stroth, S., Langmann, A., Wolff, N., Roessner, V., Roepke, S., Poustka, L., & Kamp-Becker, I. (2021). Differentiation of autism spectrum disorder and mood or anxiety disorder. Autism, 136236132110396. https://doi.org/10.1177/13623613211039673

Wright, M. N., & Ziegler, A. (2017). ranger: A Fast Implementation of Random Forests for High Dimensional Data in C++ and R. Journal of Statistical Software, 77(1). https://doi.org/10.18637/jss.v077.i01

