## Supplementary Material for "Using machine learning to improve diagnostic assessment of ASD in the light of specific differential diagnosis"

### Supplementary Methods

ASD diagnosis was performed by a multidisciplinary team (at least two trained and experienced clinicians), using all available information from ADOS, caregiver interviews, home videos, neuropsychological and IQ testing, school reports, and further differential diagnostic assessments to arrive at a best estimate clinical consensus decision (ICD-10, WHO 2014). Individuals were labeled as "ASD" if they received an ICD-10 diagnosis of F84.0, F84.1, or F84.5. Participants that still had a suspicion of ASD but no clear diagnostic decision after the assessment were excluded from the analysis.

All data processing, machine learning, statistical analysis and visualization was performed in R 4.0.3 (R Core Team, 2020), using tidymodels 0.1.3 (Kuhn, Max, 2020). Tidymodels is a collection of R packages designed to streamline model based data analysis, including machine learning methods. We used broom 0.7.6, dials 0.0.9, dplyr 1.0.5, ggplot2 3.3.3, infer 0.5.4, modeldata 0.1.0, parsnip 0.2.5, purr 0.3.4, recipes 0.1.16, rsample 0.0.9, tibble 3.1.1, tidyr 1.1.3, tune 0.1.5, workflows 0.2.2, and yardstick 0.0.8. Furthermore, the following R packages were used: doParallel 1.0.16, plotROC 2.2.1, readr 1.4.0, stringr 1.4.0, cutpointr 1.1.0, vip 0.3.2.

For parameter optimization, we used Bayesian Tuning (implemented in tune 0.1.5), i.e., an algorithm that performed an initial evaluation of a set of 25 different random parameter combinations. Iterations of further parameter combinations based on the results of the previous iterations were evaluated until the performance did not increase for at least ten iterations or reached a maximum of 50 iterations.

Model training was performed on a high performance computing cluster (Scientific Computing Cluster of the GWDG, *https://www.gwdg.de/web/guest/hpc-on-campus/scc*) , using 50 cores to train the models of the repeated cross validation cyle of the outer folds for each model. A medium partition was used, providing 4-8GB of RAM for each core at runtime.

*Model specificity.* To validate the specificity of our models we compared the overall evaluation metrics for the respective specific model (as described above) against its performance on the respective non-included participant groups (e.g., the performance of the ANX model to classify in the context of CD), see Figure S5. For unbiased results, this calculation was performed separately per outer cross-validation fold, subsequently averaged, and compared to each other using two-sample t-tests. Only those individuals were used within each fold that were not included in the respective training set.

To construct the webapp (*https://msrlab.shinyapps.io/ml_beta_app_v3/*, we used shiny 1.6.0 (implemented in R 4.0.3 and R Studio 1.4.1103) and deployed the app on shinyapps.io. For the app, four final random forest models (ASD vs. CD; ASD vs. ADHD; ASD vs. ANX; unspecific) were trained on the whole dataset. It is designed in a way, that a complete ADOS examination can be entered in less than 30 seconds on any computer with internet access. The app takes ratings of an ADOS module 3 examination and runs a prediction through one of the selected models. The prediction results in a value between 0 and 1, with higher values coding for a higher probability of an ASD diagnosis according to the random forest model. For each model, an optimal cutoff is calculated according to the training dataset (calculation of the optimal cut-points for maximized sensitivity and specificity as described in the main text). For illustration purposes, the distribution of ASD and Non-ASD samples across the model prediction values (training sample), as well as the cut-off point and the individual prediction value for the new datapoint is given (see Figure S4)

### Supplementary Results

*Model specificity.* The CD, ANX, and ADHD models performed significantly better (with respect to AUC) within their specific diagnostic sample than for the other diagnostic samples, see Figure S5. (CD-sample: CD-models better than ANX-models ($T=3.65$, $p<0.0005$), CD-models better than ADHD-models ($T=3.50$, $p<.001$; ANX-sample: ANX-models better than CD-models ($T=4.64$, $p<0.00005$), ANX-models better than ADHD-models ($T=5.39$, $p<0.00001$); ADHD-sample: ADHD-models better than ANX-models ($T=6.15$, $p<0.00001$), ADHD-models better than CD-models ($T=5.89$, $p<0.00001$). Both CD- and ANX-models also performed significantly better than the unspecific model (CD: $T=4.02$, $p<0.0003$, ANX: $T=5.78$, $p<0.000001$), and the ADHD models showed a statistical trend towards better performance in comparison to the unspecific models ($T=1.58$, $p=0.0585$ (one-sided)).

### Supplementary Tables

| ADOS items | |
|---|---|
| ANX | Anxiety |
| ARSC | Amount of Reciprocal Social Communication |
| ASK | Asks for Information |
| CONV | Conversation |
| DGES | Descriptive, Conventional, Instrumental, or Informational Gestures |
| EMO | Empathy/Comments on Other's Emotions |
| ENJ | Shared Enjoyment in Interaction |
| EXPE | Facial Expressions Directed to Examiner |
| EYE | Unusual Eye Contact |
| IECHO | Immediate Echolalia |
| IMAG | Imagination/Creativity |
| INJ | Self-Injurious Behavior |
| INS | Insight into Typical Social Situations and Relationships |
| LLNC | Language Production and Linked Nonverbal Communication |
| MAN | Hand and Finger and Other Complex Mannerisms |
| NESL | Overall Level of Non-Echoed Spoken Language |
| OACT | Overactivity |
| OINF | Offers Information |
| OQR | Overall Quality of Rapport |
| QSOV | Quality of Social Overtures |
| QSR | Quality of Social Response |
| REPT | Reporting of Events |
| RITL | Compulsions or Rituals |
| SINT | Unusual Sensory Interest in Play Material/Person |
| SPAB | Speech Abnormalities Associated with Autism |
| STER | Stereotyped/Idiosyncratic Use of Words or Phrases |
| TAN | Tantrums, Aggression, Negative or Disruptive Behavior |
| XINT | Excessive Interest in Highly Specific Topics, Objects, or Repetitive Behaviors |

**Table S1**: Items and item abbreviations of ADOS

### Supplementary Figures

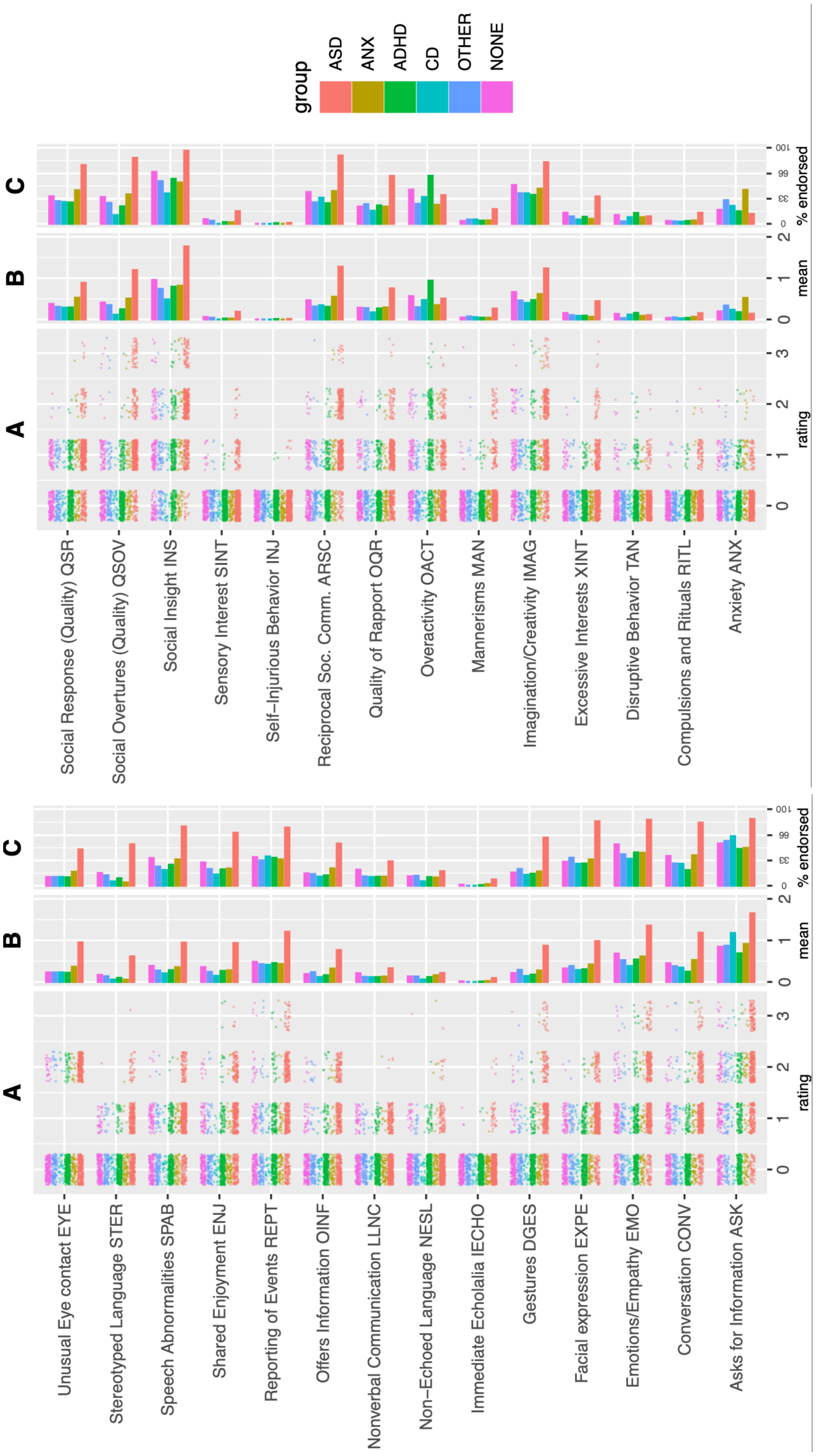


**Figure S1.** Overview of individual item ratings for the different diagnostic groups (ASD: all participants with an ASD diagnosis; ANX, ADHD, CD, OTHER, NONE: participants without an ASD diagnosis, but a specific differential diagnosis (ANX, ADHD, CD), a different ICD-F diagnosis (OTHER), or no ICD-F diagnosis (NONE). A: distribution of rating codes 0-3 for each item and diagnostic group (jittered datapoints of each single individual). B: mean score for each item, C: percentage of "symptom rating endorsed", i.e. percentage of individuals which had at least a rating of 1. Verbal description of ADOS-items are slightly abbreviated, for the full labels refer to Table S1.

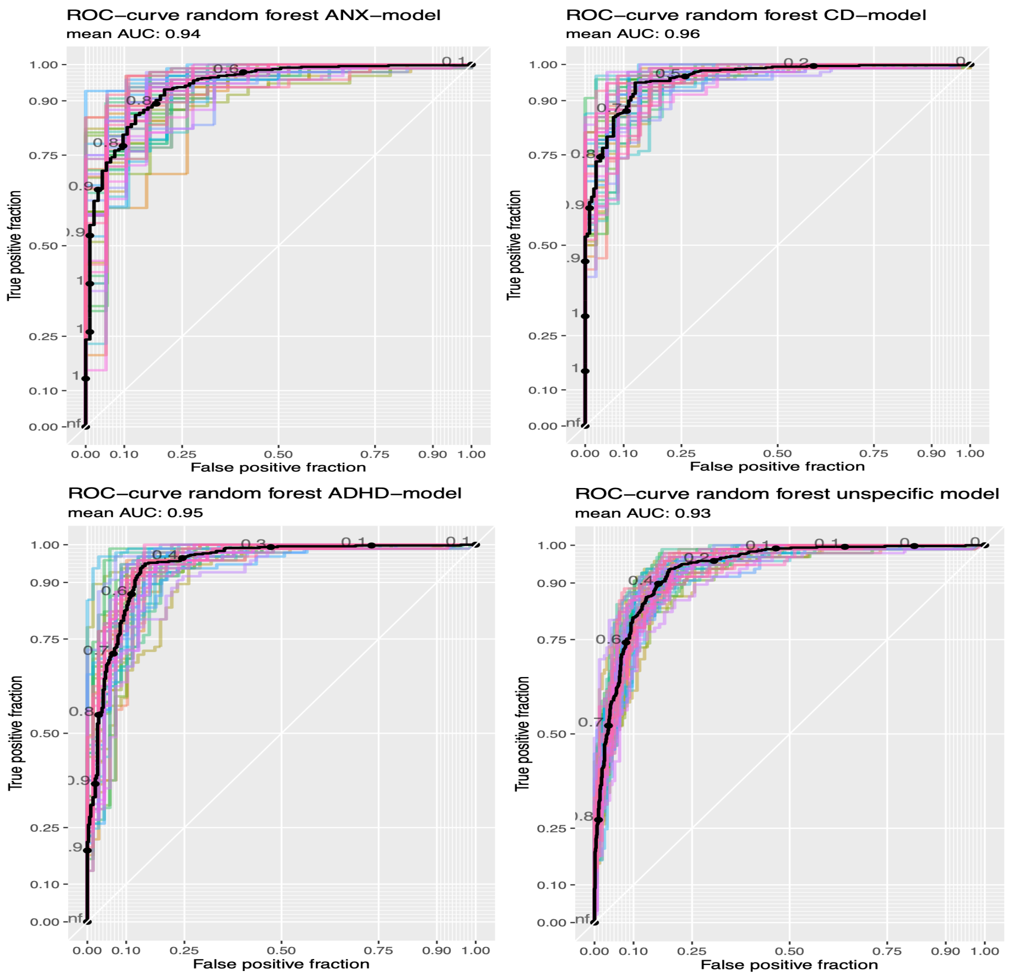


**Figure S2.** ROC (receiver operant characteristic) curves for the evaluation of the random forest models. Black line represents the ROC curve of aggregated mean values across folds. Colored lines represent ROC curves for each of the 50 evaluation models. X-axes: False positive fraction (1- specificity), Y-axes: True positive fraction (1- sensitivity)

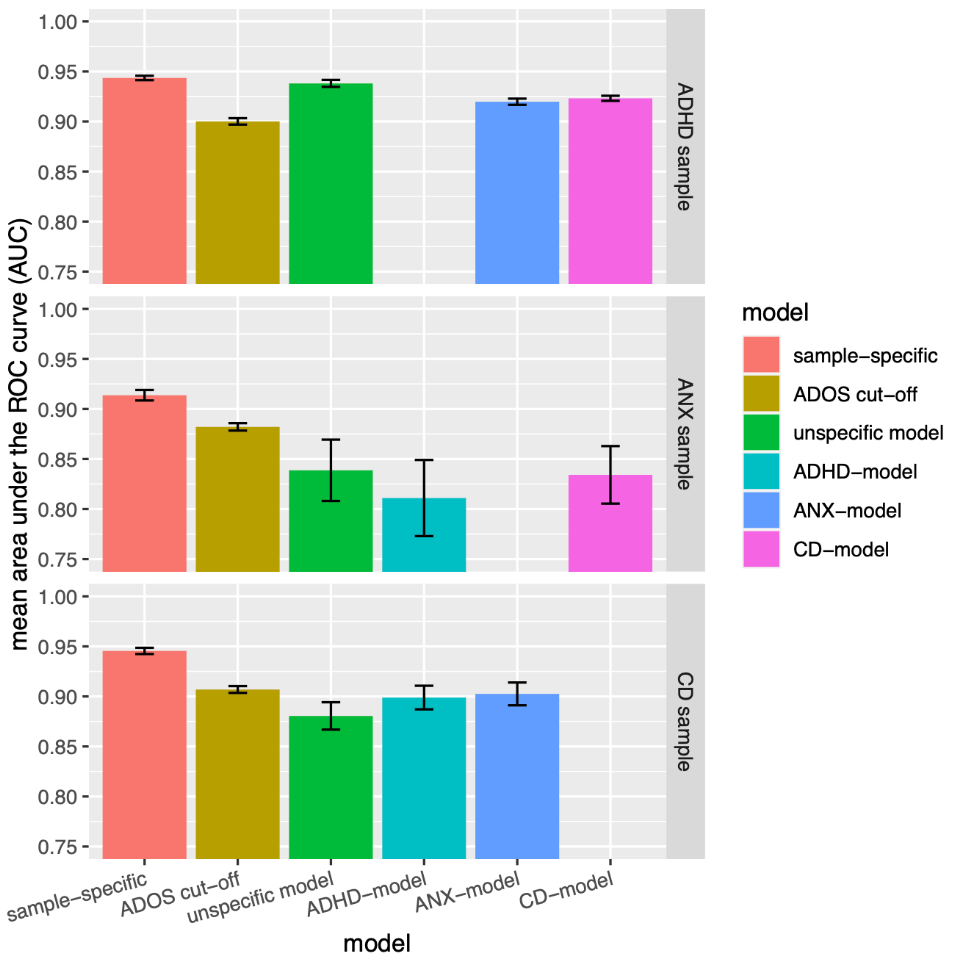


**Figure S3.** Specificity of models. Y-axis depicts the mean AUC of the group-specific model evaluation ("specific") and cross-evaluation across folds, error bars depict standard error of means. For the comparison, non-ASD cases from the other diagnostic samples were used, as well as ASD cases that had not been used for training in the respective fold.

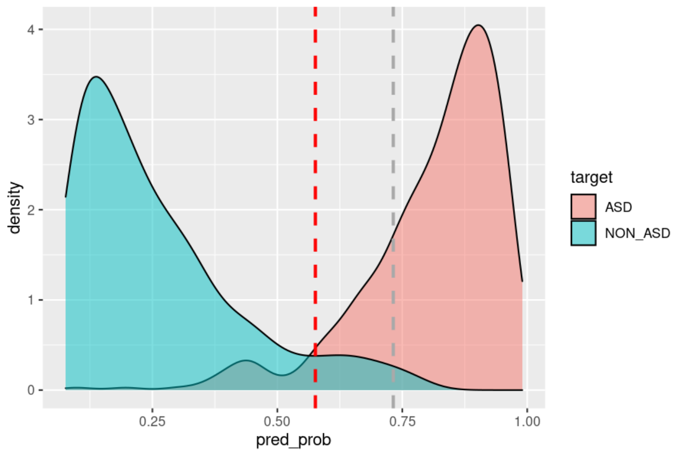


**Figure S4.** Exemplary output of the wep-app. The dashed grey line represents the individual prediction after entering a set of ADOS item scores. The dashed red line represents the optimal cut-point of the model for maximal sensitivity and specificity according to the training data. The red and cyan shaded areas represent the distribution density of model prediction values for ASD and non-ASD samples according to the training data of the respective model.
